# A Randomized Placebo Controlled Clinical Trial of a Metabolic Shifting Probiotic, Sugar Shift, for the Treatment of T2DM

**DOI:** 10.1101/2022.12.06.22283186

**Authors:** Gissel García, Josanne Soto, Lays Rodríguez, Maricela Nuez, Noraika Domínguez, Emilio F. Buchaca, Duniesky Martínez, Rolando J. Gómez, Yohanka Ávila, Martha R. Carlin, Raúl J. Cano

## Abstract

Type 2 diabetes mellitus (T2DM) is a chronic metabolic disorder characterized by hyperglycemia, insulin resistance and chronic inflammation. Probiotics have been claimed effective in the management of obesity and type 2 diabetes mellitus. BiotiQuest™ Sugar Shift is a symbiotic formulation rationally designed for the endogenous conversion of glucose and fructose to support restoration of the human gut microbiota, modulation of intestinal glucose, and the production of anti-inflammatory metabolites.

We report the results of a 12-week, double blind, placebo-controlled study designed to evaluate Sugar Shift in Cuban T2DM patients. Clinical parameters, including fasting and 2h post-prandial glucose, hemoglobin A1c, a lipid panel, insulin, creatinine, and serum lipopolysaccharide levels were assessed. Microbiome composition was assessed by 16S amplicon sequencing of the variable region V3-V4 of the 16S rRNA gene. Metabolic biomarkers were inferred from microbiome data by Kruskal-Wallis H test and LEfSe.

Fasting glucose, Insulin, and serum LPS levels decreased significantly at day 84 as compared to day 1 in the treated group and to control group. Hb A1c remained stable in the treatment group as compared to the controls but not show significant improvement in the study period.

Microbiome analysis showed significant increase in Chao1 alpha diversity in the treated group between day 1 and day 84. Taxonomic and functional biomarkers revealed significant differences between the Day 1 and Day 84 microbiome profiles in the treatment group, primarily associated with acetate, propionate, and butyrate production.

Our results indicate that Sugar Shift can be a suitable adjunct therapy to standard of care therapy in the management of T2DM based upon the improvement in key inflammatory and insulin resistance markers. These results were interpreted as an indication of favorable microbiome changes during the course of the treatment for 12 weeks.

## Introduction

Diabetes mellitus (DM), is a metabolic disease characterized by alterations in glucose metabolism (1). Four main etiological categories are described: type 1 (T1DM), type 2 (T2DM), seasonal DM and the other types of DM, which fall into the same etiological category. T2DM comprises 90% of all cases (1). Statistics from 2018 show that this metabolic condition affected 537 million individuals worldwide (https://idf.org/aboutdiabetes/what-is-diabetes/facts-figures.html) and it is estimated that by 2045, about 625 million people will suffer from this disease (2). Cuba does not escape this situation; in the Statistical Yearbook of the Ministry of Public Health (MINSAP) of 2019, a prevalence of this disease of 66.7% was reported, 2% higher than that reported in 2018 (3). Therefore, diabetes mellitus in general, and type 2 in particular, is considered an emerging pandemic disease by the international scientific community, since its prevalence increases in all countries regardless of their level of development (4).

T2DM is diagnosed when there is a state of glucose intolerance (glucose ≥ 11.1mmol/L) and fasting elevated blood sugar levels (≥ 7mmol/L or126 mg/dl) are detectable. (1, 5). Treatment requires lifestyle change, timely incorporation of oral antidiabetics, including metformin, and maintaining of glycosylated hemoglobin (HbA1c) at levels below 7%. However, in clinical practice, patients do not always achieve glycemic control (4).

It is proposed that genetic and environmental factors are associated with the development of this etiological category (1, 6). In the context of these factors, the gut microbiota plays a central role in this disease as it is able to modify the biological and metabolic functions of the organism, regulate inflammation and as is essential for the mechanisms of innate and adaptive immunity (7).

Disease severity, immunodeficiencies, obesity and insulin resistance have been associated with imbalances in the gut microbiota. This phenomenon, known as dysbiosis, occurs from multiple causes (4). These include unbalanced diets (8), environmental toxin exposures (9, 10) and the indiscriminate use of antibiotics (11, 12), among other factors that have been associated with the pathogenesis of T2DM (13–15). A diet rich in carbohydrates and processed foods favors the reduction of beneficial bacteria and the growth of others that, for example, increase the secretion of lipopolysaccharides (LPS). Increased secretion of LPS induces insulin resistance (1, 4, 7).

Recent studies support that the consumption of certain formulations of beneficial bacteria in association with the nutrients they produce could reshape the intestinal microbiota and, in this way, improve glucose levels and insulin resistance in individuals suffering from T2DM (16). This group of living microorganisms, administered to a host in appropriate forms and quantities, improves or restores the intestinal microbiota and, consequently, its metabolic and immunological functions. For this reason, these organisms have been referred to as probiotics. These mixtures are administered as a symbiotic formulation with prebiotics and non-digestible plant fibers that stimulate the growth of the probiotic organisms and other beneficial intestinal microorganisms (16, 17).

There are many commercial probiotic formulations on the market which mostly comprise a limited variety of commercial probiotic strains in wide use. However, many are not balanced formulations and there is very little strain or species diversity in the currently commercialized products. Many formulas do not contain any prebiotic at all and most do not evaluate or show the analysis of the genetic and metabolic profile of the consortium of bacteria as a working community in the formula. The BiotiQuest™ Sugar Shift (Sugar Shift) preparation, on the other hand, is a formulation designed with the aid of a community-based flux balance analysis model (18, 19), coined the BioFlux™ model, an in silico analytical platform for modeling microbial consortia. The computational modeling provides for the design of formulations with a balance of the flow of metabolic inputs and outputs over time to target a specific metabolic output and improved sustainability. The symbiotic formulation Sugar Shift consists of eight strains of bacteria with GRAS (Generally Recognized as Safe) classification. The core properties of this formula’s design are the production of mannitol, with the concomitant reduction of glucose and fructose, the production of anti-inflammatory components, such as butyrate (20) and the antioxidant reduced glutathione (21).

This study was designed to determine whether Sugar Shift would be an effective adjunct therapy to the prevailing standard of care to T2DM. This study was designed as a double blind, placebo-controlled study aimed to evaluate the supplement Sugar Shift in a population of Cuban T2DM patients during the course of 12 weeks. This decision for the venue was made based on the excellence of the National Health System in Cuba and the track record in well-conducted clinical trials (22) as well as on the reports indicating that the epidemiology of T2DM in Cuba is similar to that of the rest of the world (3).

### Hypothesis

T2DM is a chronic metabolic disorder characterized by hyperglycemia, insulin resistance and chronic inflammation (23). Several studies have reported gut microbiome dysbiosis as a factor in rapid progression of insulin resistance in T2DM that accounts for about 90% of all diabetes cases worldwide (1). Probiotics, prebiotics, symbiotics, and postbiotics benefit metabolic diseases management, especially obesity and type 2 diabetes mellitus (24). Sugar Shift is a symbiotic formulation rationally designed for the conversion of glucose and fructose to support restoration of the human gut microbiota, the modulation of intestinal glucose, and the production of anti-inflammatory metabolites and therefore stabilize blood glucose, improve insulin resistance, and reduce inflammation.

## Materials and Methods

### Study design

The study was designed and conducted as a randomized and double blind, placebo-controlled trial to investigate the effect of Sugar Shift as a supplementary therapeutic approach for T2DM with ISRCTN registry number ISRCTN48974890. The full trial protocol is provided herein as supplementary material, including the English translation of the approved document submitted to the Institutional Review Board for approval. The CONSORT flowchart of the study is shown in Fig 1.

**Fig 1.**
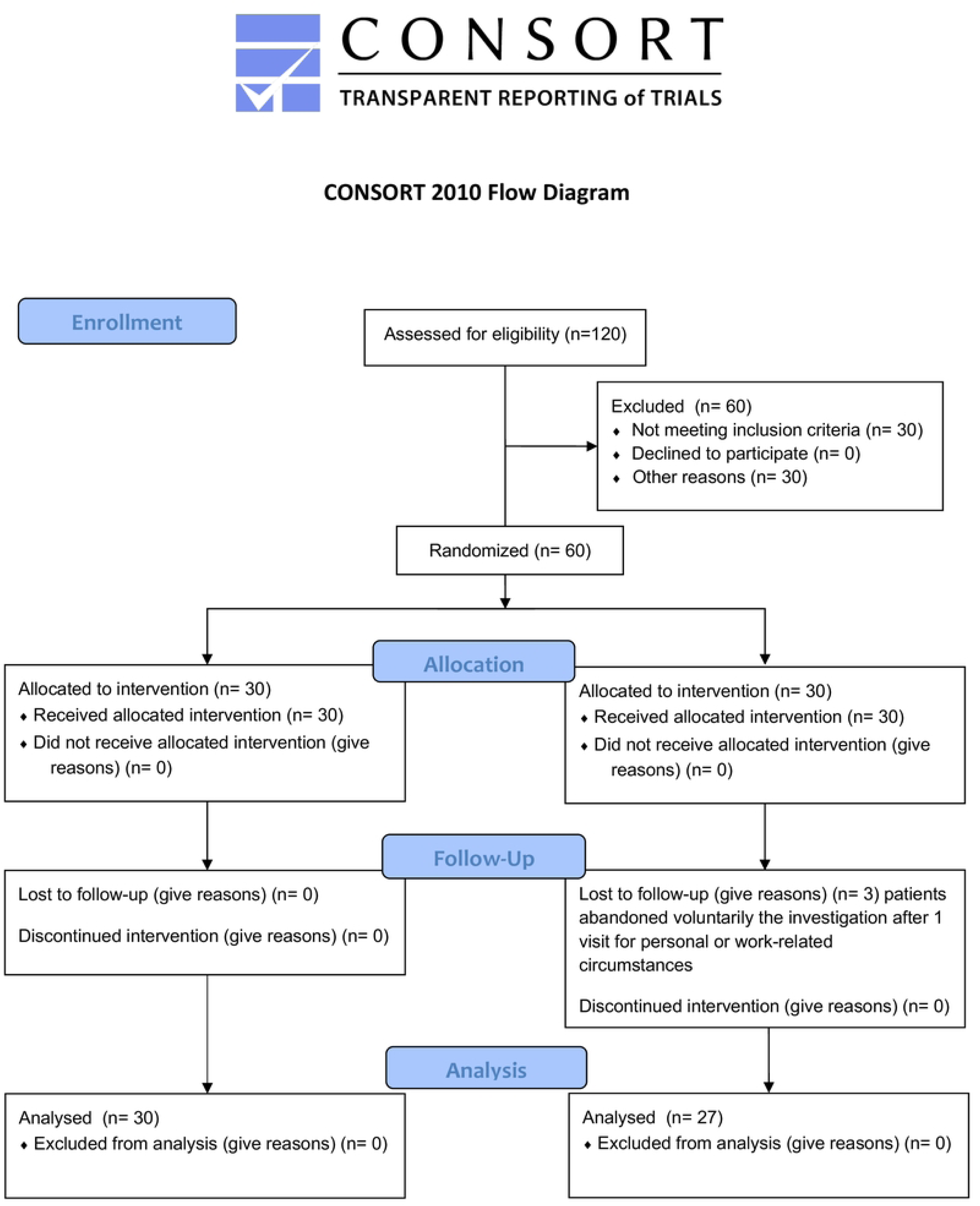
CONSORT 2010 Flowchart of the Study

The study was conducted at Hermanos Ameijeiras Clinical Hospital, an Academic Medical Center in La Habana, Cuba. Fig 2 show outlines the study design and timeline. No changes to the approved protocol by the Institutional Review Boards were made.

**Fig. 2.**
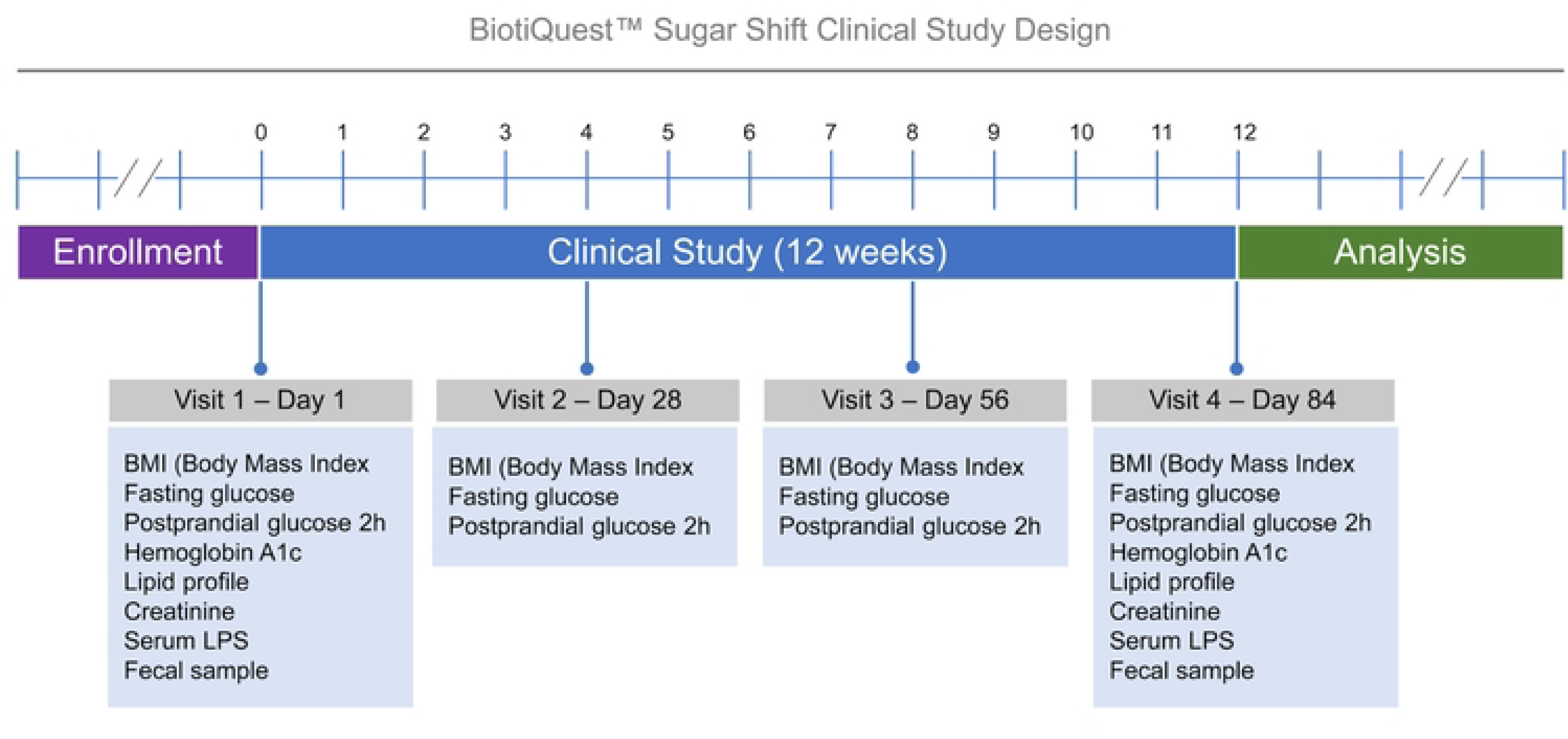
Schematic representation of the clinical study design. The diagram represents the various elements and metrics used in the present study. The Purple zone, representing the enrollment period, extended for more than 6 months, until the number of participants exceeded the minimum required to achieve the power of 0.8. The clinical study, represented by the blue bar, extended for exactly 12 weeks, while the Analysis period (green bar) extended for more than 4 weeks. The boxes contain the assays performed at the time periods indicated (in weeks) in the diagram

### Ethical Considerations

This trial was evaluated and approved under the Hermanos Ameijeiras Ethic Committee for Clinical Investigation and The Hermanos Ameijeiras Scientific Council. The Sugar Shift Probiotic supplements were also evaluated and approved for their use by the Ethical Committee at the National Institute of Nutrition of Cuba and The Cuba Ministry of Health. The study was conducted considering the Good Clinical Practice and in accordance with the code of ethics of the World Medical Association Declaration of Helsinki (25).

### Eligibility

All patients diagnosed with T2DM (26) between 30 and 65 years old and body mass index (BMI between 28-40) at baseline, who attended the diabetes specialized consultations of the Hermanos Ameijeiras Hospital and who gave their consent to participate in the study, without excluding sex or skin color, were selected. Patients with chronic kidney disease, onco-proliferative diseases and pregnant women were excluded.

### Power and sample size consideration

The sample size was estimated with a two-sided alpha of 0.05, 95% confidence level, standardized mean difference of 0.75, test power of 80%, expected proportion of losses of 10%. Based on this information, the minimum required sample size was estimated to be 28 individuals per group; adjusted for losses for various reasons, the sample size was established at 32. The sample consisted of 64 patients who consented to participate in the research.

### Patients Recruitment

The recruitment period of the patients was from June 2021 to April 2022. The period was extended due to Covid and lockdown. Patients were surveyed according to the inclusion criteria and the discretion of the attending physician for inclusion in the study, also we ask for information if the patients had had SARS-CoV-2 infection.

### Randomization

The included patients were assigned to two treatment groups by randomization technique: Group A (Sugar Shift) consisted of 32 patients who were assigned to the treatment group, Group B (control): consisted of 32 patients who were given a placebo. Assignment to treatment groups was randomized using the Epidemiological Analysis from Tabulated Data (EpiData 3.1) software (http://www.epidata.dk). The list of random numbers for each group was placed in the pharmacy. The assignment was made when the patient’s compliance with the inclusion and signature criteria for the informed consent to participate in the study was verified. The products were dispensed by a pharmacist who distributed a corresponding number of capsules to each patient, according to the order of arrival at the pharmacy and this was noted in the medical record. Both Sugar Shift and placebo were packaged in indistinguishable foil packaging as well as same size capsules to maintain blinding. Subjects may be unblinded if deemed medically necessary by their provider and the study principal investigator (PI) in Cuba. However, in this study no subjects were unblinded.

### Delimitation and operationalization of variables

The variables measured in this study were as follows: age (in years completed); sex (female, male); nutritional evaluation (according to body mass index (BMI) = Weight in Kg / Height in m^2^: low weight: < 18.5 Kg/m^2^, normal weight: 18.5-24.9 Kg/m^2^, overweight: 25-29.9 Kg/m^2^, obesity: ≥ 30 Kg/m^2^.

The Primary response variable was hemoglobin A1c (HbA1c) (%). The Secondary Response Variables were fasting glucose (FBG) (mmol/L), postprandial glucose (PPG) –2h (mmol/L), insulin (mU/mL); total cholesterol (mmol/L), triglycerides (mmol/L), high density lipoprotein (HDL-c) (mmol/L), low density lipoprotein (LDL-c) (mmol/L) and lipopolysaccharide (LPS) levels determination mml/L).

### Trial intervention

Subjects consumed Sugar Shift or placebo (Control Group) daily, two capsules per day, 12 hours apart. The foil packaging containing the capsules were distributed every 28 days over the 12-week study period after blood sample collection for clinical determination (Visit 1, Visit 28 and Visit 56 as timeline shown in Fig 1).

### Test substance

The “test substance”, Sugar Shift was manufactured by BlisterPak Pro, LLC (Lafayette, Colorado). Each capsule contains 96 mg (18 billion CFU) of a bacterial consortium of eight strains of GRAS-classified bacteria that include *Bacillus subtilis* De111™, *Bifidobacterium bifidum, Bifidobacterium longum, Lactobacillus paracasei, Lactobacillus plantarum* TBC0036, *Lactobacillus reuteri, Leuconostoc mesenteroides* TBC0037, and *Pediococcus acidilactici.* Each capsule contains 370 mg of prebiotics and fillers consisting of inulin, microcrystalline cellulose, D-mannitol, and stearic acid.

### Placebo control

The capsules, indistinguishable in appearance from the test substance, were similarly manufactured by BlisterPak Pro, LLC (Lafayette, Colorado) and contained the same ingredients in identical proportions as the test substance sans the microbial consortium. Each placebo capsule contained 50 mg each of the prebiotics inulin and D-mannitol and 270 of the fillers microcrystalline cellulose, and stearic acid.

### Organization and sample collection

The patients, once randomized, were divided into subgroups for office visits to facilitate instruction and clinical evaluation of the participants as well as sample collection. Each group was cited every 28 days for sample collection and delivery of supplements and clinical evaluation. Fecal samples from ten patients from each cohort group were collected for microbiome characterization studies at Day 1 and Day 84 of the study (for a total of 40 fecal samples).

All patients included in the study were measured for: FBG, PPG, cholesterol, triglycerides, HDLc, LDLc, HbA1c, LPS, and fasting insulin. To obtain the biological samples, the patients were instructed to fast for 8 hours. Whole blood samples were obtained by venipuncture. The serum was obtained after centrifugation of the primary sample within 1 hour to 1 hour 30 minutes after extraction, to be processed and aliquoted for preservation and subsequent use. The preservation of aliquots of fasting samples was carried out in the containers intended for this purpose at a temperature - 20 °C until used for LPS determination (Fig 1). The blood sample for the HbA1c determination was dispensed in blood collection K3EDTA tubes (Henso Medical (Hangzhou) Co., Ltd.).

To ensure data security, electronic records of subject data were maintained using a dedicated Microsoft Excel Database, Access to electronic databases was password protected and limited to study staff and clinical staff supporting the subject’s care. The Hermanos Ameijeiras Hospital managed the security and viability of the information technology infrastructure

### Adverse events (AE)

This was a minimal risk study that involved the use of a commercially available dietary supplement and AEs related to the study product beyond the potential for bloating or excessive gas if taken in excess (beyond recommended dosing) were not anticipated. Creatinine levels were measured to assess, the impact, if any, on the kidney of the participants in all study groups.

### Clinical Determinations

The determinations were made in a modular immunochemical autoanalyzer Cobas 6000, from Roche Diagnostic that meets the requirements stipulated in Directive 98/79/EC of the European Parliament and the Council of the European Union (EU) on *in vitro* diagnostic medical devices, following the Standard Operating Procedures (SOPs) in force, and with the quality standards required for them (Table 1).

**Table 1.** Quality Standards used with the Immunochemical Autoanalyzer Cobas 6000

| Paramenter | Range of Quality Control Standard |
| --- | --- |
| FBG | 4.2 - 7 mmol/L |
| PPG | ≤ 10 mmol/L |
| Cholesterol | 3.6 – 5.2 mmol/L |
| Triglycerides | 0.5 – 1.85 mmol/L |
| HDLc | ≥ 0.9 mmol/L |
| LDLc | 2.6 – 3.35 mmol/L |
| HbA1c | ≤ 6% |
| Insulin | 2.6 – 24.94 mIU/mL |
| Creatinine | 47.6 – 113.4 µmol/L |

### LPS determination

The determinations of serum LPS levels (results expressed in Endotoxin Units) of participating patients were made using the commercial kit ToxinSensor^TM^ Endotoxin Detection System (Version 11242021), following the manufacturer’s instructions (www.genscript.com).

### Microbiome Analysis

Fecal samples were collected using DNA/RNA Shield™ Fecal Collection Tubes (Zymo Research, Irvine, CA) and stored at room temperature until processed.

#### 16S amplicon sequencing

16S amplicon sequencing was performed by EzBiome (Gaithersburg, MD, USA). Genomic DNA concentration was quantified using the Qubit fluorometer (ThermoFisher, USA). The 16S rRNA V3-V4 regions within the ribosomal transcript were amplified using the primer pair (Illumina-F: and Illumina-R, which contains the gene-specific sequences and Illumina adapter overhang nucleotide sequences using EzBiome’s established protocol. It is noteworthy to indicate that microbiome reference standards (ZymoBIOMICS Microbial Community Standard, Zymo Research, Irvine, CA) were used to quality control the process.

#### Amplicon Taxonomic Assignment and Functional Prediction

Taxonomic profiling of 16S sequencing data was carried out by directly uploading forward and reverse paired end reads to the EzBioCloud microbiome taxonomy profiling platform (www.ezbiocloud.net) as described elsewhere (27). Briefly, the cloud application of the EzBioCloud detects and filter out sequences of low quality regarding read length (<80BP or >2,000bp) and averaged Q values less than 25. Denoising and extraction of non-redundant reads are carried out using DUDE-Seq software (28). The UCHIME (29) algorithm is applied against the EzBioCloud 16S chimera-free database to check and remove chimera sequencing. Taxonomic assignment is performed using the USEARCH program (30) to detect and calculate the sequence similarities of the query single-end reads against the EzBioCloud 16S database. Sequencing reads are clustered into OTUs at 97% sequence similarity using the UPARSE algorithm (31). Reads from each sample were clustered into many OTUs using the UCLUST (32) tool with the above-noted cutoff values. For the EzBioCloud 16S-based MTP pipeline, the PICRUSt2 algorithm (33) was used to estimate the functional profiles of the microbiome identified using 16S rRNA sequencing. The raw sequencing reads were computed using the EzBioCloud 16S microbiome pipeline with default parameters and discriminating reads that were encountered in the reference database. The functional abundance profiles of the microbiome are annotated based on bioinformatics analyses, specifically by multiplying the vector of gene counts for each OTU by the abundance of that OTU in each sample, using the KEGG (Kyoto Encyclopedia of Genes and Genomes) (34) orthology and pathway database.

#### Amplicon Comparative Statistical and Bioinformatic Analyses

Subsampling, generation of taxonomy plots/tables and rarefaction curves, and calculation of species richness, coverage, and alpha and beta diversity indices were carried out in EzBioCloud Application. Briefly, microbial richness was measured by ACE, Chao1, Jackknife and the number of OTUs found in the microbiome taxonomic profile (MTP) index. The Shannon, Simpson and Phylogenetic α-diversity indices were applied to estimate the diversity for each group using the Wilcoxon rank-sum test (35). Beta diversity was calculated with Bray-Curtis (36), UniFrac and Generalized UniFrac (37) distances based on the taxonomic abundance profiles. Permutational multivariate analysis of variance (PERMANOVA) (38) were applied to measure the statistical significances of β-diversity. Different groups were clustered with principal component analysis (PCoA) based on abundance Jaccard distance metric. Kruskal-Wallis H test (39) and LEfSe (40, 41) analysis were performed to determine enrichment in the assigned taxonomic and functional profiles between groups. Taxonomic levels with LEfSe values higher than 2 at a p-value < 0.05 were considered statistically significant. The ggplot2 package in the R program (version 3.4.3., R Foundation for Statistical Computing, Vienna, Austria) were used to visualize the LEfSe differences between the groups. All the calculated p values were two-tailed and considered statistically significant at p<0.05.

### Statistical analysis

#### Clinical data

The information collected was organized in an Excel database. These data were then exported to the Statistical Package for Social Sciences (SPSS) version 23.0 system for analysis and R For Windows - R: The R Project For Statistical Computing, version 4.2.0. Samples were characterized through the description of the variables of interest. Qualitative variables were summarized in absolute numbers and percentages.

Quantitative variables were summarized in mean and standard deviation when data presented a normal distribution, verified by the Kolmogorov-Smirnov test for normality (42); if they did not present normal distribution, they were summarized in median and interquartile range (IR).

To detect differences between the groups according to qualitative variables, the chi-square test (χ^2^) (43) was used. For the same purpose, the Student’s t-test (44) was applied in the case of age (quantitative variable).

The comparison of medians of the variables that provided information on the effect of therapeutics between the different times (day 1, day 28, day 56 and day 84), was performed using the Friedman test (45). In the case of the initial (day 1) and final (day 84) comparison, the Wilcoxon signed range test (46) was used.

The comparison of variance between both groups that provide information about of the effect of probiotics therapeutics between the different times (day 1, day 28, day 56 and day 84), was performed using the F-Test Two-Sample for Variances (47). Analysis of covariance (ANCOVA) is a general lineal model which blends ANOVA and regression. ANCOVA evaluates whether the means of a dependent variable (DV) are equal across levels of a categorical independent variable (IV) often called a treatment, while statistically controlling for the effect of other continuous variables that are not of primary interest, known as covariates (CV) or nuisance variables. Mathematically, ANCOVA decomposes the variance in the DV into variance explained by the categorical IV, and residual variance. Intuitively, ANCOVA can be thought of as ^..^ adjusting^..^ the DV by the group means of the CV (s) The ANCOVA model assumes a linear relationship between the response (DV) and covariate. (CV) (48). We worked with the level of significance α = 0.05 in all hypothesis tests.

To detect differences according to quantitative LPS and insulin determinations between the groups and in the groups respect day 1 to day 84 the paired sample t-test was applied as well as Pearson’s correlation (43). Also, the analysis assuming unequal variance was made to detect the differences between day 1 and day 84 at a level of significance α = 0.05 to test the hypothesis.

#### Microbiome data

All statistical analyses were performed using packages ‘vegan’ v2.5-6, ‘phyloseq’, ‘ggpubr’ and ‘ggplot2’ v3.3.2 in R 3.6.3 (https://www.r-project.org/. For microbiome analysis, rarefaction depth was set at 25,000 reads. Shannon diversity index. (49), Chao1 Index (50) and Good’s Coverage of library (51) were used to evaluate alpha (within sample) diversity. Beta (between sample) diversity was examined using multidimensional scaling analysis (MDA) (52) of Bray-Curtis (36) and Jaccard (53) distances. The Wilcoxon rank-sum test (35) was used to compare alpha diversity values between groups (p > 0.05). Statistical significance of beta-diversity distances between stool processing workflows was assessed using PERMANOVA (38) with 999 permutations. Alpha diversity group significance was calculated using nonparametric Kruskal-Wallis H test (39). Taxonomic features descriptive of each cohort were identified using Linear Discriminate Analysis (LDA) Effect Size (LEfSe) (40), employing the Kruskal-Wallis (KW) sum-rank test (39) to detect features with significant differential abundance with respect to the class of interest. The biological significance is then investigated using a set of pairwise tests among subclasses using the (unpaired) Wilcoxon rank-sum test (35). As a last step, LEfSe uses Linear Discriminant Analysis(54) to estimate the effect size of each differentially abundant feature and, if desired by the investigator, to perform dimension reduction. Benjamini and Hochberg false discovery rate correction factor (55) errors in null hypothesis testing when conducting multiple comparisons.

## Results

### Demographic and clinical characteristics

Demographic and clinical characteristics at the beginning of the study were similar in both randomized groups. A total of 60 individuals were included for this investigation, 30 per group, but 3 patients abandoned voluntarily the investigation after visit 1 for personal or work-related circumstances. All of them were in the Control Group. Ultimately, the study was comprised of 57 patients, 30 in the Study Group (Sugar Shift Group) and 27 in the Control Group (Control Group).

Among the patients studied, there was a preponderance of the female sex for both groups (Sugar Shift Group 60.0%, Control Group 51.9%). On average, the patients were approximately 55 years old, minimum age 33 and maximum 65. No significant differences were found in terms of sex (p = 0.725) or age (p = 0.120).

The distribution of patients according to nutritional assessment showed overweight and obesity (Sugar Shift Group 53.3% and Control Group 51.9%). Patients were less commonly of normal weight. No significant differences (p = 0.722) were detected in both groups.

Regarding the type of treatment, diet plus oral hypoglycemic agents prevailed in both cohorts (Sugar Shift Group: 63.3%, Control Group: 55.6%). No significant differences were found regarding the type of treatment between the two groups (p = 0.549).

### Safety of the Symbiotic Product

No adverse events were reported by participants in the Sugar Shift group. One participant in the Control Group complained of bloating during the second visit (day 28).

Creatinine levels were measured and monitored to assess the impact, if any, Sugar Shift has on kidney function (56). Normal levels of serum creatinine are 65.4 µmol/L to 119.3 µmol/L (57).Mean creatinine levels were not significantly different (p = 0.24) between the Control Group at day 1 (77.17 µmol/L) and the Sugar Shift Group at day 1 (77.98 µmol/L). Similarly, there were no significant differences (p = 0.21) between the control and the study group at the conclusion of the study with values of 77.17 µmol/L in the control group and 77.97 µmol/L in the group receiving Sugar Shift.

### Clinical Chemistry

#### Fasting Glucose (FBG)

Fasting glucose levels showed that in three months this parameter is stabilized in the Sugar Shift Group as compared with the Control Group See Fig 3.

**Fig 3.**
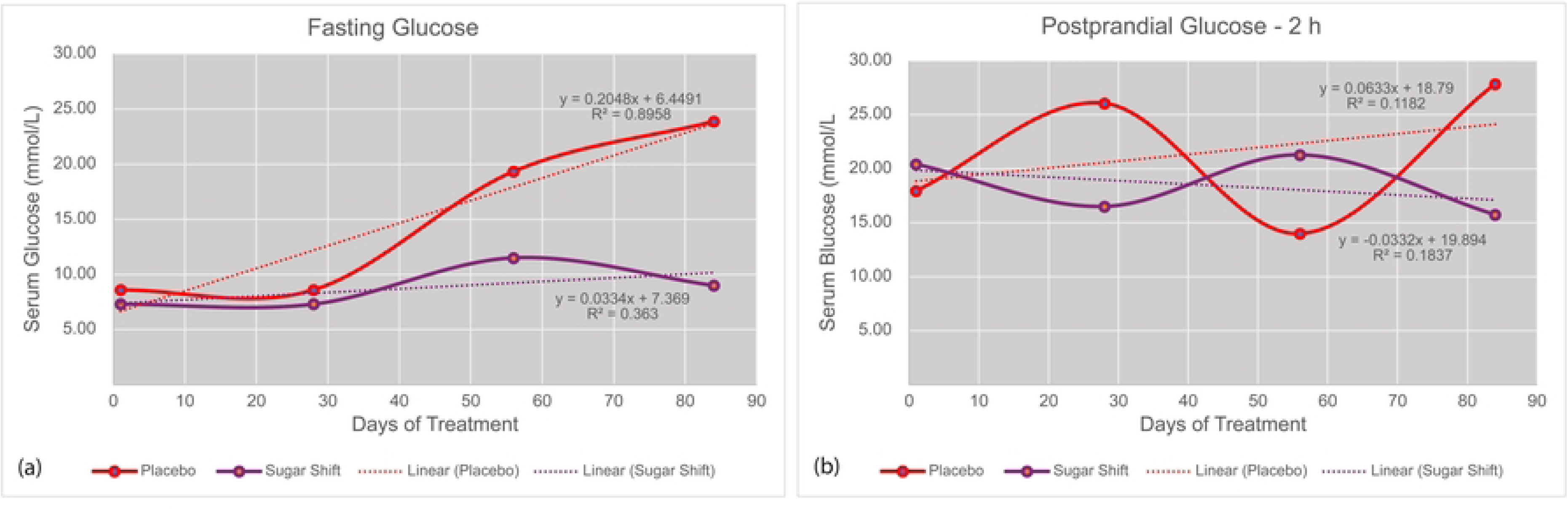
Variance trends in the Control Group and Sugar Shift Group cohorts during the 84-day study period. Variances for each sample set was calculated and plotted at four sample periods indicated in Fig 2. Trend lines for each cohort are represented by dashed line. Solid lines represent the plot of the actual variances. Red-colored lines represent results for the Control group and Purple-colored lines represent the variances of the Sugar Shift group. (a) Variance trends for fasting glucose. (b) Variance trends for 2-hour postprandial glucose

In the Sugar Shift Group, the median FBG was 7.5 mmol/L (135 mg/dL) at the beginning of the study and remained at 7.2 mmol/L at the second (day 28) and third doses (day 56) and resulted in an FBG value of 7.2 mmol/L (129.6 mg/dL) at the end of treatment, with no significant differences between different times (p = 0.942). In the Control Group, the behavior was different. There was a significant (p = 0.001) increase in the median FBG value between the first and last day of treatment (7.5 mmol/L vs. 8.0 mmol/L) (Table 2).

**Table 2.** Levels of T2DM biomarkers during the course of the study.

| Variables | Cohort | Study Days |  |  |  | P<br>value |
| --- | --- | --- | --- | --- | --- | --- |
|  |  | Day 1 | Day28 | Day 56 | Day 84 |  |
|  |  | Descriptive statistics: Median (IR)* |  |  |  |  |
| Fasting<br>Glucose<br>(mmol/L) | SS** Group | 7.5† (4.5) | 7.7 (5) | 7.4 (5.1) | 7.2 (4.1) | 0.942<sub>a</sub> |
|  | C** Group | 7.5 (3.9) | 7.3 (4.3) | 7.8 (4.1) | 8.0 (2.6) | 0.001<sub>a</sub> |
| 2h Post-Prandial<br>(mmol/L) | Sugar Shift<br>Group | 11.1 (7.3) | 11.5 (5.9) | 10.1<br>(8.4) | 10.9 (8.2) | 0.646<sub>a</sub> |
|  | C Group | 9.5 (5.8) | 10.1 (6.1) | 9.3 (6.3) | 10.8 (6.3) | 0.013<sub>a</sub> |
| HbA1C<br>(%) | SS Group | 7.2 (1.9) | ND | ND | 7.2 (1.9) | 0.262<sub>b</sub> |
|  | C Group | 6.8 (2.1) | ND | ND | 7.2 (3.4) | 0.387<sub>b</sub> |
| HOMA IR<br>index | SS Group | 8.65 (5.5) | ND | ND | 7.12 (3.9) | 0.002 |
|  | C Group | 11.54<br>(14.2) | ND | ND | 12.51 (8.2) | 0.191 |

\* IR = Interquartile Range
\*\*SS = Sugar Shift treated cohort; \*C = Placebo control cohort
a= Friedman Test
† Multiply the value in mmol/L by 18 to obtain values in mg/dL

In consideration of the differences observed at the end of treatment between both groups (day 56 and Day 84), we performed the analysis of variance between both groups. The variability of FBG in Control Group is much greater (23.88) than in Sugar Shift Group (8.26) after 12 weeks of treatment (84 days) (Table 3, Fig 3a). The Analysis of Covariance (ANCOVA) (48) was in concordance between both groups at 84 days. The ANCOVA results show a negative covariance between groups (z=-6.85, p(z)= 7.62257E-12), indicating that fasting glucose increases in the Control group and decreases in Sugar Shift Group at the end of 12 weeks (Table 3).

**Table 3:** Comparison of variances between groups in fasting glucose levels

| Variable |  | Study Days |  |  |  |
| --- | --- | --- | --- | --- | --- |
|  |  | Day 1 | Day28 | Day 56 | Day 84 |
|  |  | Descriptive statistics: Variance (Mean) |  |  |  |
| Fasting Glucose (mmol/L) | Sugar Shift Group | 7.30(8.29) | 7.31 (8.29) | 11.49 (8.48) | 8.99(8.31) |
|  | C. Group | 8.59(8.43) | 8.59(8.43) | 19.34(8.34) | 23.9 (9.4) |
| P(F<=f) one-tail |  | 0.33 | 0.33 | 0.089 | **0.006** |
| P(Z<=z) one-tail |  | 0 |  |  | 3.81128E-12 |
| P(Z<=z) two-tail |  | 0 |  |  | 7.62257E-12 |
| F Critical one-tail |  | 1.891466319 |  |  |  |
| z Critical one-tail |  | 1.644853627 |  |  |  |
| z Critical two-tail |  | 1.959963985 |  |  |  |

#### Postprandial glucose – 2h

Regarding postprandial glucose, the median values in Sugar Shift Group were 11.1 and 11.5 mmol/L in the early stages of the study and in the last two measurements, showed a slight decrease (10.1 and 10.9 mmol/L) that was not significant (p = 0.646). In the Control Group, however, a slight increase at the end of the treatment was significant with respect to the stable values of the Sugar Shift group (p = 0.013) (Table 2). The declining trend in pp glucose from day 1 to day 84 was similar to FBG (Fig 3b) even when no significant differences was observed at the end of the study in analysis of variance between both groups (p = 0.62). The ANCOVA results show a negative covariance between the Control group (26.817) and the Sugar Shift group (−0.0289), indicating that postprandial glucose increases in the Control group and decreases in Sugar Shift Group at the end of 12 weeks. The kinetics of cohort variances throughout the study for both FBG and PPG can be observed in Fig 3.

#### Glycosylated Hemoglobin (HbA1C)

The analysis of HbA1C showed that no significant change occurred in either the Sugar Shift Group or the Control Group. Glycosylated hemoglobin was found, on average, at 7.2% in the two measurements made (day 1 and day 84) in the study group. In patients in the Control Group, at baseline, it was 7.3% and, at the end, 7.4%. No significant differences were found (Table 2). It is noteworthy that a 90-day trial period may not have been sufficient for either group to be a clear indicator of improvement in view that the RBC cycle is approximately 120 days (58).

#### Cholesterol

The data collected during this aspect of the study, as well as the metadata of the participating patients, are listed in Table S1 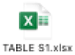. Mean cholesterol levels were not significantly different (p =0.371) in the Sugar Shift Group while there was a significant (p = 0.0007) increase in mean cholesterol values in the Control Group. The F-test Two-Sample for Variance analysis revealed a significant increase (p = 0.00032) in variance from day 1 to day 84 for the Control Group but not significant (p = 0.391) for the Sugar Shift Group. Mean cholesterol values for the Control Group were 4.41 mmol/L at day 1 and 5.14 mmol/L at day 84. For the Sugar Shift Group, mean cholesterol values remained steady at 4.31 mmol/L at day 1 and 4.42 mmol/L at day 84. Variances for this latter group were 0.645 and 0.584, respectively. For the Control Group at days 1 and 84, the variances were 0.311 and 1.644, respectively.

#### HDL and LDL Cholesterol

The means of the HDLc values of day 1 and day 84 for the Sugar Shift group were 1.19 and 1.39, respectably. Similarly, the mean values for the Control group were 1.17 and 1.12 for Days 1 and 84, respectively. These values were not statistically significant when the two groups were compared. The variances, however, between Day 1 and 84 of the SS group were 0.12 and 2.72, respectively. These were statistically significant (p = 0.00012). The Control group, however showed no significant (p = 0.137) variances between Day 1 (variance = 0.07) and Day 84 (variance = 0.11).

#### Triglycerides

Triglyceride levels were reduced, on average, from 2.1 to 1.7 mmol/L in the Sugar Shift Group and from 2.4 to 1.6 mmol/L in the Control Group, results that were significant in both cases (Sugar Shift Group: p = 0.005; Control Group: p = 0.002). The analysis of variance between groups no showed significant differences.

#### Insulin

Insulin levels were measured for all study participants and then analyzed statistically using both the t-Test (paired) and F test for analysis of variance. The Control Group (Days 1 and 84) and Sugar Shift Group (Day 1) participants showed higher insulin concentrations than those in the Sugar Shift Group after 84 days of treatment (Table 4). Using the t-Test Paired Two-Sample for Means the p values for Sugar Shift Group Day 1 and Sugar Shift Group Day 84 was p = 0.02 and interpreted as significant.

**Table 4.** Statistical analysis using the t-Test for Paired Samples for Means for serum insulin in the treated and control groups

| Metric | t-Test: Paired Two Sample for Means - Insulin (mIU/L) |  |  |  |  |  |  |  |
| --- | --- | --- | --- | --- | --- | --- | --- | --- |
|  | *SS_D1\** | *SS_D84* | *C_D1\** | *C_D84* | *CD_D1\** | *SS_D1* | *C_D1\** | *SS_D84* |
| Mean | 9.00694744 | 7.08535518 | 11.2669246 | 12.334936 | 11.2669246 | 7.86968928 | 12.334936 | 6.2201627 |
| Variance | 92.241692 | 29.057633 | 131.74252 | 172.73747 | 131.742529 | 37.454623 | 172.737474 | 15.9212268 |
| Observations | 31 |  | 27 |  | 27 |  | 27 |  |
| Pearson Correlation (59) | 0.913198142 |  | 0.936299149 |  | -0.126860439 |  | -0.047453887 |  |
| df | 30 |  | 26 |  | 26 |  | 26 |  |
| t Stat | 2.068872866 |  | -1.183398856 |  | 1.290814199 |  | 2.283333364 |  |
| P(T<=t) one-tail | 0.023634798 |  | 0.123679404 |  | 0.104064286 |  | 0.015413179 |  |
| t Critical one-tail | 1.697260887 |  | 1.70561792 |  | 1.70561792 |  | 1.70561792 |  |
| P(T<=t) two-tail | 0.047269596 |  | 0.247358807 |  | 0.208128571 |  | 0.030826357 |  |
| t Critical two-tail | 2.042272456 |  | 2.055529439 |  | 2.055529439 |  | 2.055529439 |  |

\*SS = Sugar Shift Cohort, C = Control Cohort, D1 = Day 1 of the study; D84 = Day 84 of the Study.

Conversely, the Control group (Day 1 and Day 84), and Control Day 1 and Sugar Shift Group Day 1 were 0.12 and 0. 11, respectively. However, the p value. For this latter pair, the Mean for Day 1 participants was 9.01 with a Variance of 92.24 as compared to the Sugar Shift Group Day 84 group, which showed a Mean of 7.09 and a Variance of 29.05. By way of comparison, the Mean values for Control Group Day 1 and Control Group Day 84 were 11.27 and 12.34, respectively with corresponding Variances of 131.44 and 170.74.

The F-test showed similar results. Day 1 of both groups were not significantly different (p = 0.192) while for Day 84, the Control and Sugar Shift participants were significantly different (p = 2.41 x 10^-6^) with means of 12.5 µU/L for the Control Group and 8.64 µU/L for the Sugar-Shift treated group, representing a 30.9% reduction in insulin levels in those patients receiving the symbiotic formulation.

#### Insulin Resistance

HOMA-IR, as an indicator of insulin resistance, was calculated according to the formula: fasting insulin (µU/L) x fasting glucose (nmol/L)/22.5. The Control Group (Days 1 and 84) and Sugar Shift Group (Day 1) participants showed higher HOMA-IR values than those in the Sugar Shift Group after 84 days of treatment (Fig 4). Boxplots were generated using the script “*ggboxplot*” in the library “*ggppubr” (*https://cran.r-project.org/web/packages/ggpubr/index.html) with “jitter” to show data distribution.

**Fig 4.**
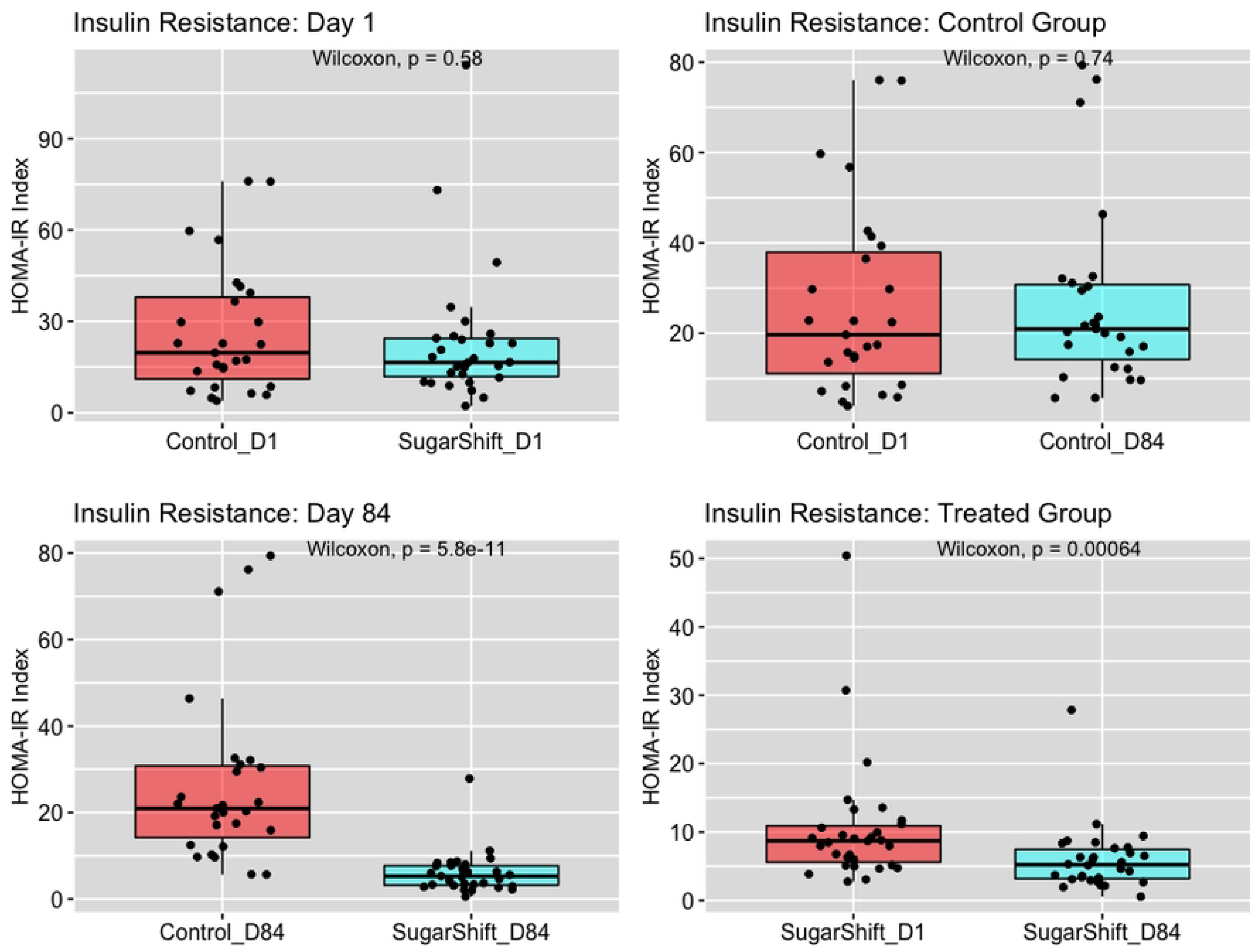
Boxplots showing distribution data gathered from HOMA-IR calculations of before and after treatment for both the Control Group- and Sugar Shift Group-treated cohorts. Boxplots were plotted with <u>ggboxplot</u> using pairwise comparisons among the four possible groups. The “jitter” function was added to show the distribution of datapoints for each of the pairwise comparisons and to assess the relationship between the measurement variable and the categorical variable. The significance (p = 0.05) in Chao1 indices between each group pair is illustrated as the probability value as calculated using the Wilcoxon signed-rank test (values on top of each graph)

Using the F-Test Two-Sample for Variance, the p values between the Control Group (Day 1 and Day 84) and Control Group Day 1 and Sugar Shift Group Day 1 were 0.24 and 0.18, respectively. However, the p value for Sugar Shift Group Day 1 and Sugar Shift Group Day 84 was p = 0.0071 and interpreted as significant. For this latter pair, the Mean HOMA-IR index was 11.27 with a Variance of 94.19 as compared to the Sugar Shift Group Day 84 group, which showed a Mean of 8.84 and a Variance of By way of comparison, the Mean values for Control Group Day 1 and Control Day 84 were 11.27 and 12.33, respectively with corresponding Variances of 131.74 and 172.74.

#### Serum Lipopolysaccharides (LPS)

Serum LPS was measured for all participants in the two cohort groups. The results are summarized in Fig 5 and Table 5. There was a significant difference between day 1 and day 84 in the Sugar Shift Group (0.00099). Similarly, significant differences were observed in the levels of serum LPS between the Control and Treated groups after 84 days of treatment (p = 0.012). No significant difference was noted in the Control Group (p = 0.0.922).

**Fig 5.**
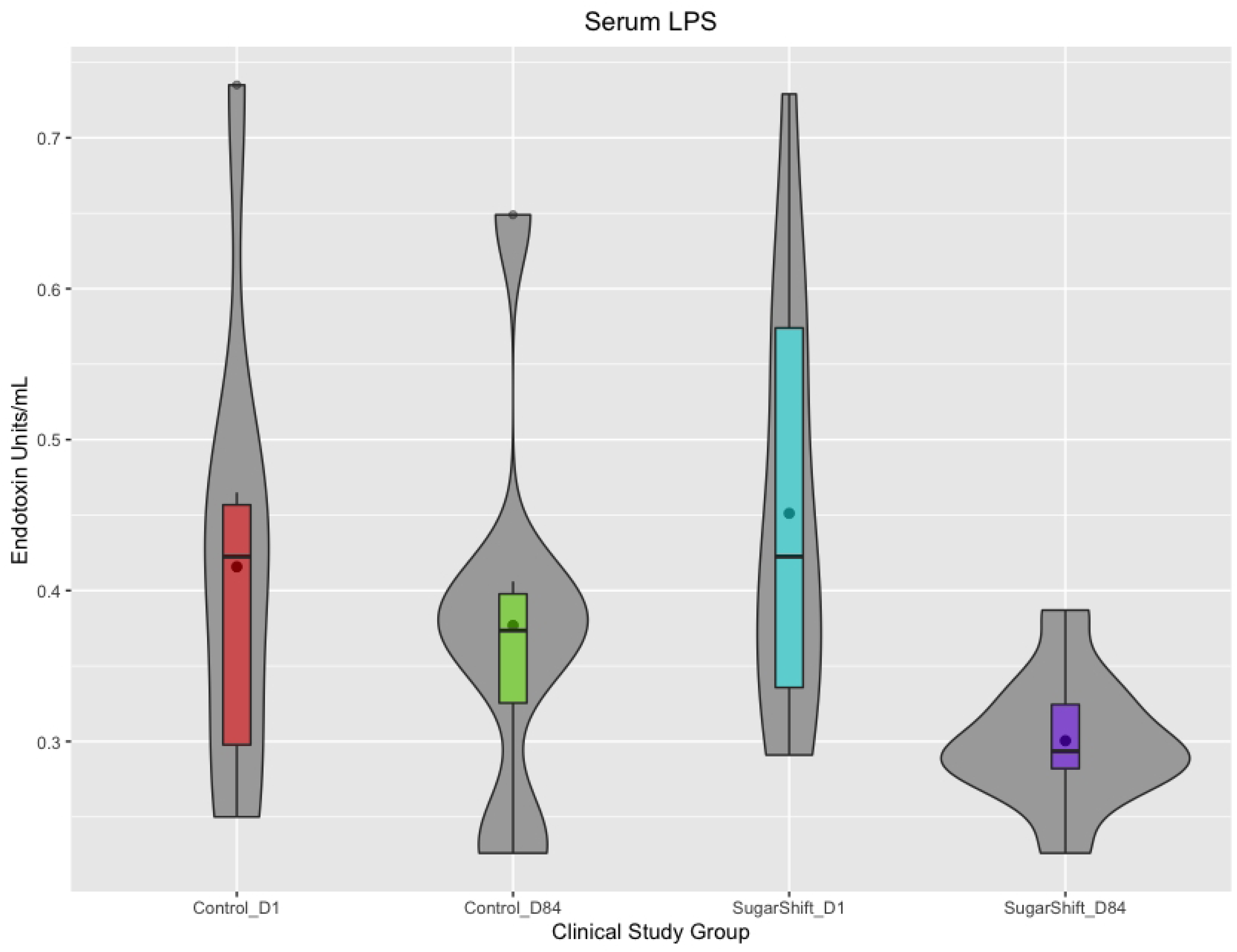
Violin plots showing the distribution of LPS values for each study group. Violin plots were chosen to show the distribution of LPS values of samples within each study group. Wider sections of the violin plot represent a higher probability that members of the population will take on the given value; the skinnier sections represent a lower probability. The colored dots within the bar plot represent the mean values for the study group while the black line indicates the median.

**Table 5.**
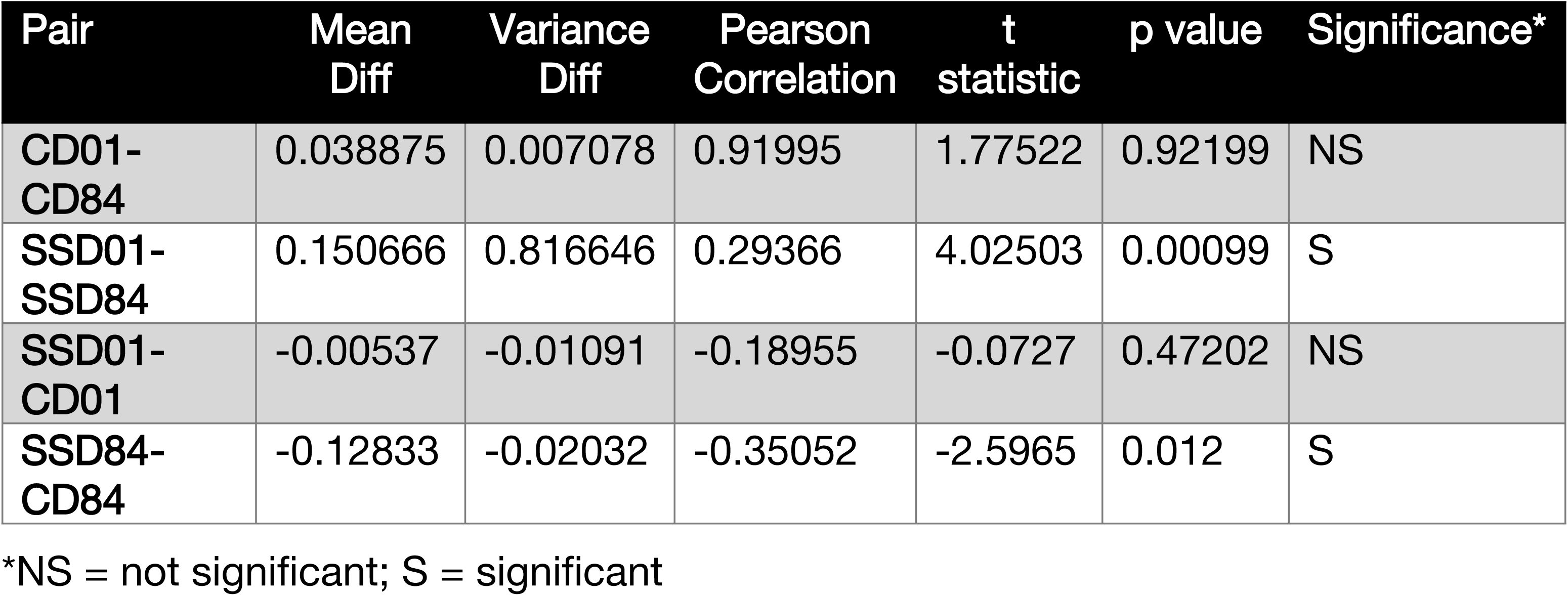
Paired samples t- Test for means among cohorts and sample times for serum LPS

### Microbiome analysis

A total of 40 samples, 10 samples from each cohort were collected before and after treatment and processed as described above. The mean sequence length per sample was 21,333 ± 7,288 base pairs (bp) with a mean Q value of 37.2 as determined using FastQC (60).

#### Alpha Diversity

Species richness, as determined using the Chao1 Index (50), showed a significant increase (Wilcoxon’s signed-rank test) (61) between Day 1 of treatment with Sugar Shift and day 84 (p = 0.0089). Similarly, there was a significant difference between the Control and the Treatment group at the end of the 12-week study (p = 0.011). No such significant differences were noted between the two groups at the beginning of the study (p = 0.80) or the Control Group at the beginning and end of the study (p = 0.80). No significant difference in the Shannon Diversity Index as measured by the Wilcoxon’s rank test was observed among the various groups. Good’s Coverage (51) of data showed no significant difference in coverage among the two groups, before and after treatment. The mean coverage was 99.83 ± 0.54. Alpha Diversity (Chao1) boxplots with “jitter” are shown in Fig 6.

**Fig 6.**
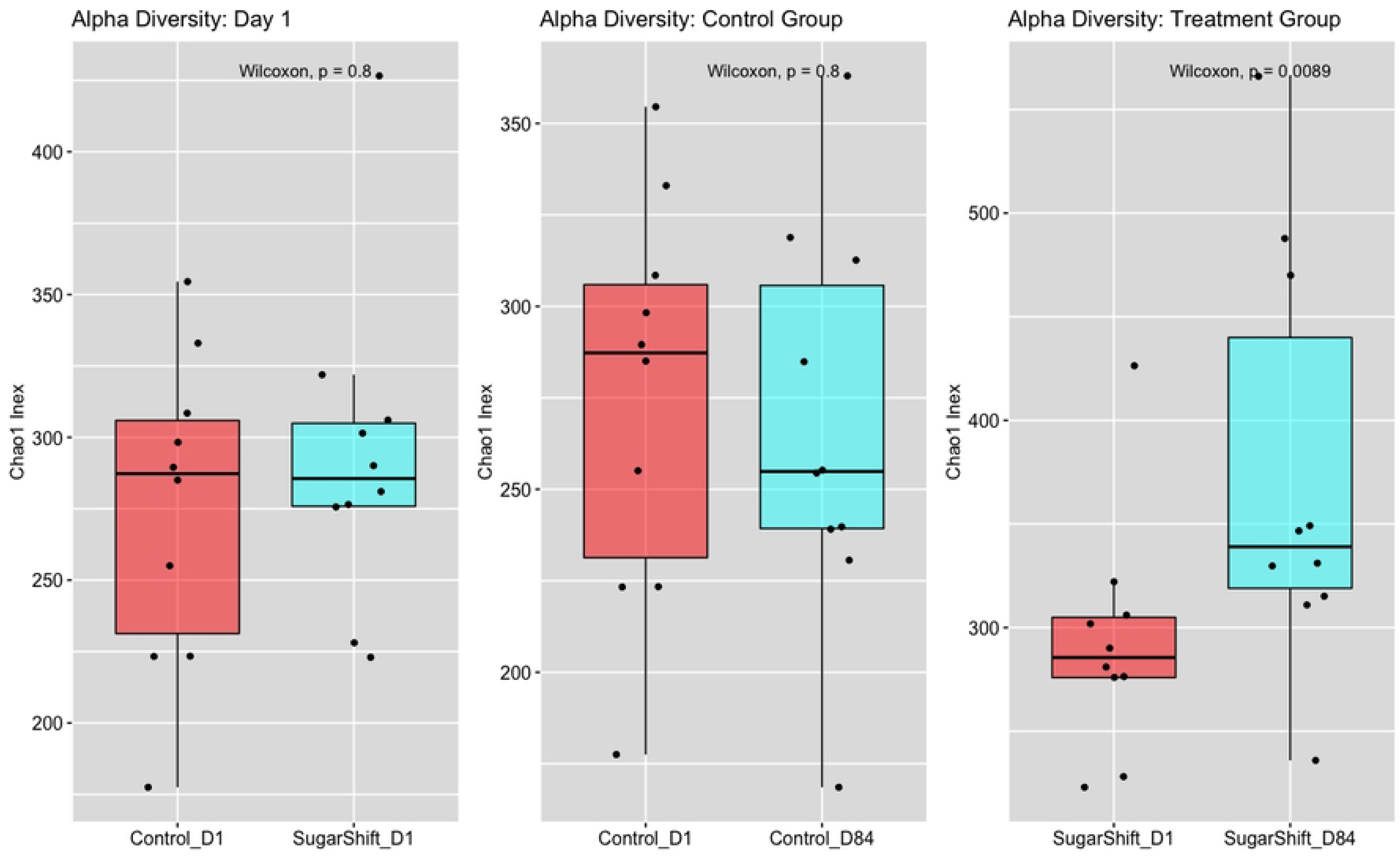
The Chao1 alpha diversity of Control Group- and Sugar Shift Group-treated patients is represented in boxplots with jitter showing inter- and intra-group data distribution. The number of species in each of the pairwise comparisons of fecal microbiomes are illustrated in these graphs as the Chao1 Index. Each dot represents a data point for each sample. The significance (p = 0.05) in Chao1 indices between each group pair is illustrated as the probability value as calculated using the Wilcoxon signed-rank test (values on top of each graph).

#### Beta Diversity

No significant changes were noted in Beta Diversity among the groups evaluated. UPGMA clustering(62) revealed that microbiome samples from individuals clustered together, regardless of the date of sample collection. While the results show a difference in Unifrac distances (37) between Day 1 and Day 84 in the Sugar Shift cohort, this difference was not statistically significant (p = 0.989).

#### Taxonomic Biomarkers

Taxonomic biomarkers were determined using both Kruskal-Wallis H test (39) and LefSe analysis (40). Two taxonomic biomarkers were identified. When comparing Day 1 and Day 84 of the Sugar Shift treated cohort, several taxonomic biomarkers were identified, both, via the Kruskal-Wallis H test and LEfSe analysis. Significant increases in abundance of several genera of the *Lachnospiraceae* family (p = 0.008), specifically *Blautia* (p = 0.02008)*, Dorea* (p = 0.02448)*, Coprococcus* (p = 0.0322) and *Kineothrix* (p = 0.2378) as well as *Prevotella* (p = 0.013) and *Bifidobacterium* (p = 0.011) and *Faecalibacterium prausnitzii* (p = 0.0018. Significant decreases in relative abundance of *Enterococcus* (p = 0.04696), *Vagococcus* (p = 0.00335), *Ruminococcus* (p = 0.04116) and *Peptostreptococcaceae* (p = 0.04508).

*Clostridium innocuum* group, showing a decrease from 0.05211 on Day 1 of the study to 0.00183 at Day 84 with an LDA effect size of 3.10536 and a p-value of 0.03051. Conversely an increase in *Agathobaculum* sp. was noted from day 1 (0.16824) to Day 84 (0.44657) with an LDA effect size of 2.34567 and a p-value of 0.4254.

#### Functional Biomarkers

Profiling phylogenetic marker genes, such as the 16S rRNA gene, is a key tool for studies of microbial communities but does not provide direct evidence of a community’s functional capabilities. PICRuSt was developed for predictive functional capabilities using 16S rRNA. Functional prediction using PICRUSt (63, 64) and LEfSe (41) revealed significant differences between the Day 1 and Day 84 of treatment with Sugar Shift in the gut microbiome in the relative abundance of microbial genes related to certain metabolic pathways associated with carbohydrate and lipid metabolism (Table 6).

**Table 6.**
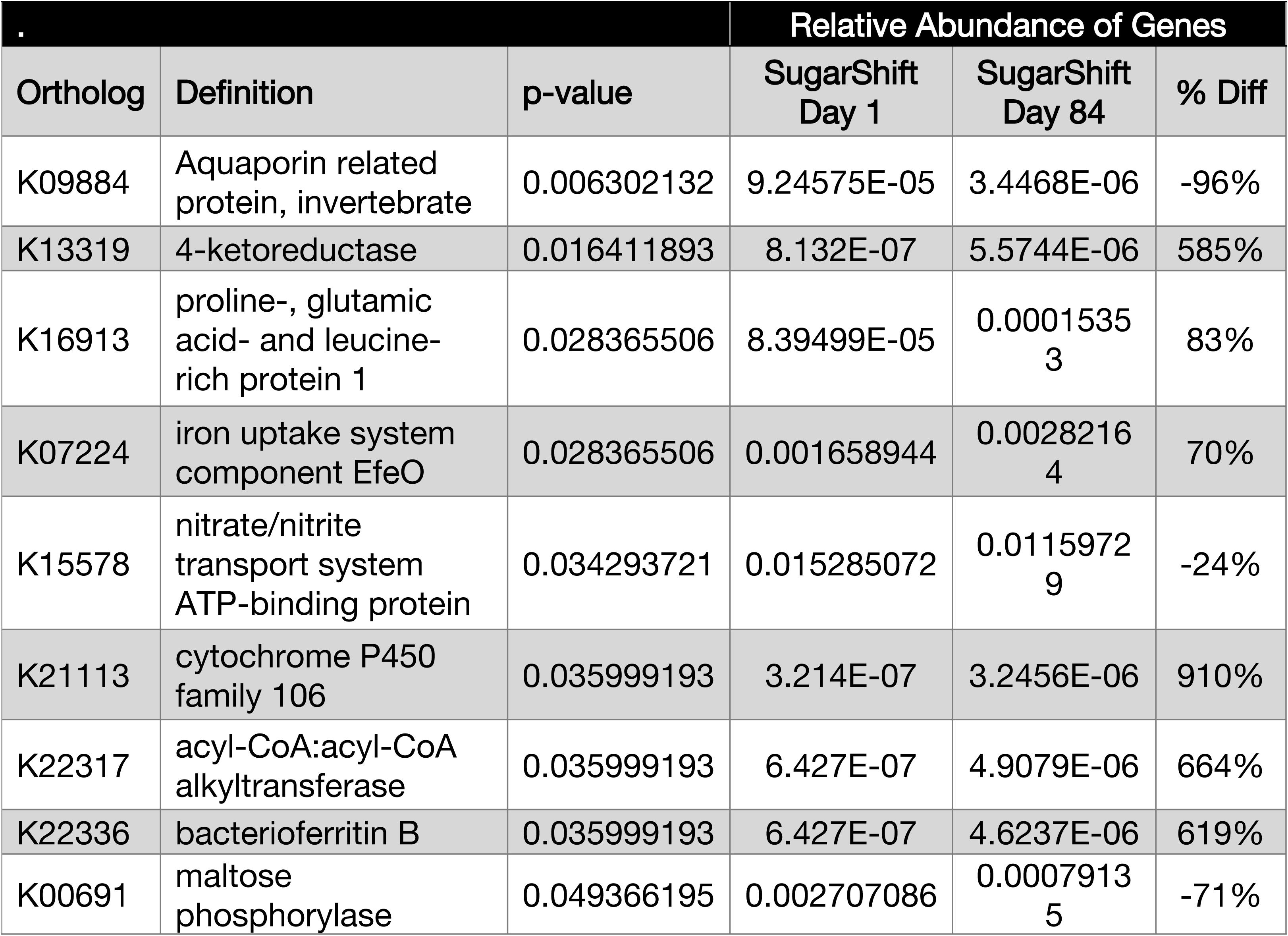
Principal Functional Biomarkers identified by PiCRUSt and LEfSe Analysis of Treated in the Sugar Shift cohort.

## Discussion

In this study, we have shown that in a short period of administration (twice daily for 12 weeks) of the symbiotic formulation Sugar Shift to T2DM patients, there were significant differences between the treated and control group on several key metrics: fasting blood glucose, insulin, HOMA-IR and serum LPS. This study also provides evidence that supports the benefits of the administration of this formulation as an adjunct therapy for the control of insulin resistance and in the lipidic profile in diabetic. The HbA1C and postprandial glucose, however, did not show significant differences as compared to placebo.

Meta-analysis studies have been summarized for a great diversity of published studies in which different probiotic formulations have been evaluated for patients suffering from T2DM (65, 66). Many of these studies have shown significant variations in the values of HbA1c and fasting glucose. (67–69). Others, however, have not shown such movements. (70, 71).

Measuring HbA1c is an accepted and valued method to estimate long-term average glycemia (72). These levels are associated with erythrocyte (RBC) longevity in peripheral circulation. Erythrocyte survival curves turnover may explain the discrepancy in the reported results (67–69, 71). The strong correlation of glycation rate with whole blood HbA1c in the DM and NDM subjects provides evidence that the whole blood determination depends on the integration of glycation rate throughout the red cell life span (73). In addition to RBC life span, there may be other variables that either attenuate or amplify the discordances between Hb A1c and average plasma glucose. In healthy adults, complete turnover average 120 days, during which time approximately 1.7 x 10^11^ cells are renewed each day (74). The analysis conducted on RBC survival curves from Cohen et al. (73) suggests an average erythrocyte circulation time of 80 ± 11 days. In view of this observation, it can be assumed that a large proportion of the glycosylated erythrocytes present in peripheral circulation of both the study cohorts at the beginning of the study were still present after 84 days. These observations may explain the similarity in the levels of HbA1c at both sampling times. A 3-month study is not sufficiently long to clearly assess the effect of the test substance (Sugar Shift) on HbA1c levels. Therefore, it is recommended that future studies may be extended for an additional 3 months to assess the impact of Sugar Shift, if any, on HbA1c levels.

### Clinical Chemistry

To analyze the effects of Sugar Shift on glycemic control evaluated by FBG, postprandial glucose and HbA1c levels, it is important to analyze the characteristics of the patients who participated in the study. As can be seen from the median values, both groups consisted of individuals with stable values of their glucose and HbA1c parameters due to previously implemented diabetic treatments. This reflects the action of the hypoglycemic treatments that most of both cohorts were taking prior to the beginning of the study as well as the intervention of the endocrinologists in their follow-up during the time of the COVID 19 pandemic, which preceded the study (75–77).

#### Serum glucose and HbA1c

At the latter part of the study period (days 56 and 84) it was observed that the FBG values of the individuals in the control group increased significantly (Fig 2). Glucose levels in the Sugar Shift Group, on the other hand, remain constant, with a tendency to decrease towards the final day of the study (day 84). These data differ significantly between study groups. F-test data showed a p = 0.006 when comparing the variance between the Control and the Sugar Shift Group at the end of the study. Covariance studies (78) showed a negative covariance (−0.6086977), that is, they are moving in opposite directions (78). These results indicate that while levels of FBG are increasing the Control Group, they are decreasing in the Sugar Shift Group, and for the 84-day study, these values remained stable in the Sugar Shift cohort.

The meta-analysis by Hu Y-m, et al. (66) on the effects of different probiotic supplements in patients with T2DM showed that, in many studies the probiotic supplements evaluated, regardless of the study design, the duration of treatment (6-12 weeks) and the doses of the supplements produce a slight reduction in FBG and an inappreciable change in HbA1c. These results are in concordance with the results presented here.

Among the possible mechanisms of probiotic therapy is promotion of a nonimmunologic gut defense barrier, normalization of increased intestinal permeability and improved gut microecology play key roles. Another possible mechanism of probiotic therapy is improvement of the intestine’s immunologic barrier, particularly through intestinal immunoglobulin A responses and alleviation of intestinal inflammatory responses, which produce a gut-stabilizing effect (79, 80).

Our results also suggest that if stabilization occurs after day 56, this trend should be evaluated over a period longer than 3 months. Studies evaluating the action of probiotics in longer periods of time find significant decrease in HBA1c and FBG values (65, 81, 82).

Another important aspect in the optimal T2DM management is the PP Glucose control. This parameter is often difficult to achieve because different mechanisms are involved. The macronutrients composition and also the sequence and timing of a meal gastric emptying/intestinal glucose absorption, gastrointestinal hormones, hyperglycemia mass action effects, insulin/glucagon secretion/action, de novo lipogenesis and glucose disposal are integrated via the central nervous system for optimal regulation and potentially, to reduce postprandial glucose fluctuations and thereby, HbA1c (83, 84). The contribution of the microbiome to this control, particularly in post prandial glucose is not well recognize yet, but our results showed a coincident tendency in the behavior of FBG and Post Prandial Glucose, so we believe that the symbiotic intervention in a period more than 12 weeks is a promising approach to regulate via gut healthy bacterial the hyperglycemia after meals in T2DM patients (85).

Considering the above, as well as the results obtained, we can hypothesize that Sugar Shift promotes a stabilization of the gut microbiome as a precursor to a noticeable decrease in FBG. This process could be due to the impact of Sugar Shift on the relative abundance and composition of the gut microbiota during the course of treatment. Sugar Shift was designed to endogenously convert glucose and fructose to mannitol in the GI tract The principal producers of mannitol in the consortium are *Leuconostoc mesenteroides and Lactobacillus reuteri* (results not shown here), and the principal glucose consumer are *B. longum* and *B. bifidum* (82). In addition, the symbiotic formulation controls the inflammation by butyrate production as indicated by an increase in the number of genes and species associated with butyrate production in the study group at Day 84. *Pediococcus acidilactici, Lactobacillus paracasei, Bifidobacterium longum* and *Lactobacillus reuteri* are the principal butyrate producers in the Sugar Shift consortium. The net production of butyrate was predicted by the BioFlux™ model to be 4.5 x 10^5^ mmol/h (86). Thus, the Sugar Shift therapy may help stabilize the gut microbial environment in 12 weeks of treatment and thereby prevent the generation of inflammatory mediators such a LPS and T2DM dysregulation (16, 80, 87).

#### Lipids

The analysis of the lipid profile shows a stabilization in the normal values of cholesterol in the Sugar Shift Group and a significant increase of these values in Control Group. Longer term studies are needed to assess the full impact on blood cholesterol. The colonization of the intestine by *Bifidobacterium* and *Lactobacillus* probiotics may affect the regulatory mechanism of conversion of cholesterol into bile acids and its elimination in the feces (88). These probiotic bacteria likely incorporate the cholesterol into their plasma membrane, convert it into coprostanol and deconjugated bile acids by the activity of the enzyme bile salt hydrolase (BSH) (89, 90). The activity of the BSH increase with the long-term colonization of the gut by *Bifidobacterium*- and *Lactobacillus*-containing probiotics, promoting a higher degree of BSH activity, thus, increasing the production of deconjugated bile acid (88). The decrease in cholesterol by this metabolic pathway would explain the slight increase in HDLc in the Sugar Shift Group, since the elimination of excess cholesterol would not be through the main pathway.

The decrease in triglyceride levels was significant in both groups. In the treated group it could be due in part to the regulation exerted by probiotic bacteria (91) linked to the diet (92) and increased physical activity of individuals in both study groups (93). The analysis of physical activity, as measured by questionnaire, showed that physical activity was higher in the Control Group compared to Sugar Shift Group (66.6% vs 50%, respectively). This suggest that the effect of the probiotics regulatory mechanism could be increase by the incorporation of physical exercise.

Animal studies indicate that the composition of gut microbiota may be involved in the progression of insulin resistance to type 2 diabetes (94). Several meta-analysis studies on the use of probiotics in T2DM show that most of these nutritional supplements decrease insulin in a short period of time significantly, even if other parameters such as HbA1c are not modified (65, 66, 91, 95). This is consistent with what was found in our study.

#### Insulin

Insulin resistance is a pathological state in which tissues do not respond normally to insulin in the process of glucose metabolism (96). Various factors have been implicated in the pathogenesis of insulin resistance (23, 97–99), including genetic predisposition, aging, obesity, and a sedentary lifestyle. More recently, the gut microbiota has been considered to be a key factor leading to the insulin resistance (100). The exact microbial mediators and mechanisms that link features of different gut microbial communities to changes in insulin secretion are not yet known (101). It is possible that microbes or their components such as muropeptides and LPS, penetrate the gut barrier and interact with receptors within the pancreas or insulin-responsive tissues and compromise endocrine control of metabolism and promote hepatic and peripheral insulin resistance, thereby worsening blood glucose. The LPSs interaction with Toll-like receptor 4 promotes inflammation in metabolic tissue and this innate immune response could be involve in the poor control of blood glucose. So, probiotics may be a promising approach to improve insulin sensitivity by favorably modifying the composition of the gut microbial community by the modification of the gut bacterial and reducing intestinal endotoxin concentrations, thus reducing inflammatory signaling and decreasing the insulin resistance (101).

### Microbiome Analysis

Species richness showed a significant increase (Wilcoxon’s rank-sum test) between Day 1 of treatment with Sugar Shift and day 84 (p = 0.0089). This suggests that the microbial consortium promotes an increased diversity in the gut microbiome. higher microbiome α diversity, along with more butyrate-producing gut bacteria, was associated with less T2DM and with lower insulin resistance among individuals without diabetes (102, 103).

Our study indicates that the use of symbiotic formulation Sugar Shift induces a significant modification of LPS concentration that is in concordance with the modification of insulin resistance, lipids modification and FBG reduction. The microbiome analysis confirms this hypothesis. The gut microbiome dysbiosis of patients with T2DM correlates well with increased LPS levels and its impact on the presence of inflammatory biomarkers in serum, which may provide insight into potential diagnostic and therapeutic approaches in the treatment of T2DM (104).

#### Taxonomic biomarkers

Gut microbiota-derived LPS is an important factor involved in the onset and progression of inflammation and metabolic diseases, including T2DM (105). The composition of the gut microbiota is altered in T2DM with a concomitant reduction of SCFA producers, notably butyrate (106). LPS, bacterial surface glycolipids, produced by Gram-negative bacteria are known to induce acute inflammatory reactions (107), particularly in the context of sepsis. However, LPS can also trigger chronic inflammation (104). In the case of T2DM and other metabolic diseases originating from chronic inflammation of the gut, the source of LPS is not an external infection, but rather an increase in endogenous production, which is usually sustained by the gut microbiota (107).

The microbiome analysis showed a significant increase (p > 0.01) in abundance of several genera of the Lachnospiraceae family in treated group as compared to control group. The compiled data from the Human Microbiome Project (108, 109) and Meta Hit (110) revealed that the human microbiota comprises 12 different phyla, of which 93.5% belong to Firmicutes, Bacteroidetes, Proteobacteria, and Actinobacteria. Among these, Firmicutes and Bacteroidetes dominate the gut microbiota in healthy subjects. The Lachnospiraceae family is a phylogenetically and morphologically heterogeneous taxon belonging to the clostridial cluster XIVa of the phylum Firmicutes (37). Lachnospiraceae belong to the core of gut microbiota, colonizing the intestinal lumen from birth and increasing, in terms of species richness and their relative abundances during the host’s life. However although, members of Lachnospiraceae are among the main producers of SCFA, different taxa of Lachnospiraceae are also associated with different intra- and extraintestinal diseases, that is why this genera of bacteria have a controversial role in health and disease (111). This association depends on the predominant species of this genera in the microbiome (111, 112).

In a normal human physiologic state, 90–95% of the SCFAs present in the colon and in the general systemic circulation are constituted by acetate, propionate and butyrate, with intraluminal concentrations of approximately 60% C2, 25% C3 and 15% C4 (113). Butyrate is a SCFA reported to be the major source of nutrition for colonic epithelial cells (114) with anti-inflammatory effects by induction of regulatory T cells, downregulation of pro-inflammatory cytokines and the Toll-like receptor (TLR) 4 receptors, thus reinforcing insulin sensitivity and glucose metabolism.(115). On the other hand, butyrate induces the Activation of G protein-coupled receptor (GPR) 43. This binding suppresses colonic inflammation, therefore protecting the liver and down-regulating insulin signal transduction in adipose tissue. Butyrate is also involved in modulating appetite and lipolysis inhibition, which in turn, decrease circulating lipid plasma levels and body weight (113). Recent studies on the composition and functionality of the microbiome in obese children demonstrated that insulin resistance was associated with decreased microbial α-diversity measures and abundance of genes related to the corresponding metabolic pathways (116).

In our results we found a significant increase of α diversity at Day 84 in the group of treatment with Sugar Shift. These data support our hypothesis because of the significant decrease in LPS concentration and in HOMA-IR Index in the treated group with Sugar Shift are related to de increase of α diversity in the microbiome of this people.

There was a significant (p = 0.025) increase in the relative abundance of *Bifidobacterium* spp. between both pre-treatment groups (0.63 ± 1.14) and the Sugar Shift Group after 12 weeks of treatment (2.75 ± 3.84). Increased relative abundances of this genus is notably reported in association with successful treatment with probiotics and in non-diabetic, “healthy” individuals (117, 118). It is also plausible to hypothesize that the Bifidobacteria present in Sugar Shift colonize the intestines and help restore the homeostasis of the gut microbiota and increase the relative abundance of SCFA producers, thus, alleviating inflammation. This interaction, however, would need to be supported by whole genome and metabolomic studies.

It is noteworthy that *Faecalibacterium prausnitzii* is a highly represented (44% increase from the same patients at the beginning of the study) in the gut microbe of Sugar Shift-treated patients. This taxon is generally abundant in the microbiome of “healthy” individuals, but it is present at reduced levels in individuals with gastrointestinal inflammatory diseases (e.g., diabetes). It has therefore been suggested to constitute a marker of a healthy gut and is associated with anti-inflammatory properties (119, 120). Among its attributes, *F. prausnitzii*, is an acetate consumer that produces butyrate and bioactive anti-inflammatory molecules such as shikimic and salicylic acids (121). This protective system SCFAs that are significant end-products that re involved in the fermentation process of the bacteria that exist in the human colon and ameliorates the effects of LPS (122). The significant decrease in serum LPS noted among the Sugar Shift-treated study group (Fig 3) may be attributed, at least in part, to the role of *F. prausnitzii* in the production of SSFAs and other anti-inflammatory substances.

#### Functional biomarkers

Putative, functional biomarkers were identified using PICRUSt2 to infer functional gene relative abundances from taxonomic data and LEfSe to determine discriminatory features that explain functional and taxonomic between the study groups. While these tools serve to identify differences based on gene relative abundance, rather than by function, it is noteworthy that several of the discriminatory orthologs identified play a role in T2DM. Many of which are related to iron metabolism, in particular orthologs K07224, K22317, and K22336. The blood sugar levels are closely related to the iron contents in islet b-cells. However, because of the higher expression levels of iron transport proteins in the islet b-cells when compared with other tissue cells, the islet b-cells are more likely to accumulate iron. Excessive levels of iron might induce excessive oxidative stress promote islet b-cell apoptosis, and ultimately affect insulin secretion and increase the risk of insulin resistance (123, 124). Circulating levels of key soluble proteins for body iron homeostasis are altered in individuals with obesity, insulin resistance and/or type 2 diabetes mellitus (125). It appears that the intestinal iron availability affects the gut microbiome, which in turns plays a central role in the absorption of iron in the intestine and bacteria-derived metabolites. This two-way interaction between iron homeostasis and the gut microbiota affects systemic glucose metabolism (126). The observation that important iron-processing orthologs are significantly overexpressed in the Sugar Shift-treated group seems to suggest that this formulation alleviates insulin resistance by promoting iron homeostasis via promotion of a “healthier” microbiome.

The high relative abundance of ortholog gene K22317 (acyl-CoA:acyl-CoA alkyl transferase) in Sugar Shift-treated individuals is noteworthy as this gene is involved in the regulation of lipid metabolism and subsequent lipid transformation.(127). This process was discussed above in relation to cholesterol levels in Sugar Shift-treated individuals.

## Conclusions

Sugar Shift is a symbiotic formulation consisting of a sustainable bacterial guild that promotes the conversion of glucose and fructose to mannitol and promotes the production SSFAs acetate and butyrate. The results indicated that consumption of Sugar Shift twice daily for 12 weeks, in conjunction with standard of medical care for T2DM (72) improves biomarkers for T2DM, including, HOMA-IR index and serum Insulin and LPS levels.

1. Fasting glucose and HbA1c levels were stabilized during the course of the study, showing a progressive improvement over the course of the study.
2. In this study it was shown that supplemental treatment of T2DM with Sugar Shift increases alpha diversity of the gut microbiome, which has been associated with a reduction of the biomarkers of T2DM (128, 129).
3. Insulin resistance was significantly improved in patients treated with Sugar Shift as compared to the Placebo group. The mechanism hypothesized included the modification of the gut microbiota with the concomitant reduction of serum LPS.
4. Inflammation biomarkers, specifically serum LPS was decreased. It was hypothesized that an increase in the butyrate-producing bacteria and Bifidobacteria were instrumental in the reduction of the inflammatory biomarkers.
5. Based on the presumed mode of action, that is reducing inflammation by lowering LPS and increasing production of butyrate, it is anticipated that this formulation might be effective in relieving the signs and symptoms of other inflammatory-based intestinal dysbioses.

In conclusion, Sugar Shift in this study, was shown to be a suitable nutritional supplement for the control of T2DM and the reduction of biomarkers associated with this dysbiosis

## Data Availability

The data underlying the results presented in the study are available from Martha Carlin.

## Acknowledgments

We would like to express our gratitude to Dr. Miguel H. Estévez del Toro, Director of the Hermanos Ameijeiras Hospital and to Yurailis Reyes, Eng. For their support in initiating and conducting the study.

This work was supported by The BioCollective, LLC.

